# Immunoglobulin Replacement Therapy in Chronic Lymphocytic Leukaemia patients with hypogammaglobulinaemia and infection; analysis of total national utilisation data in Australia 2008-2013

**DOI:** 10.1101/2021.12.29.21268534

**Authors:** Anastazia Keegan, Peta M. Dennington, Nina Dhondy, Stephen P. Mulligan

**Author notes:** Author for Correspondence: Professor Stephen Mulligan, RNSH landline, preferred contact, RNSH Dept Haematology, Secondary. The authors have no conflict of interest to declare.

## Abstract

**Objectives:** To analyse total national utilisation of immunoglobulin (Ig) replacement therapy (IgRT) for Chronic Lymphocytic Leukaemia patients with acquired hypogammaglobulinaemia and severe and/or recurrent bacterial infections.

**Methods:** In 2007, the National Blood Authority first published *Criteria for the clinical use of intravenous immunoglobulin in Australia*. The Australian Red Cross Lifeblood assessed, approved, and recorded all supply with patient demographics, distribution data, intravenous Ig (IVIg) volumes and treatment episodes. IVIg was the sole product used in Australia from 2008-2013 inclusive.

**Results:** From 2008-2013 across Australia, 2,734 individual CLL patients received 48,870 treatment episodes using a total 1,324,926 grams of IVIg therapy. Six IVIg products were available, with domestically manufactured Intragam®P accounting for 89.7% of supply. The average age for first dose was 74 years. Males received 60.6% of the total treatment episodes representing 20% more than females. The average pre-treatment IgG level was 4.03g/L±2.03g/L (range 0.30-10.50g/L). A sustained average annual increased IVIg utilisation of 5.5% was observed. There was significant regional variation consistent with differences in prescriber preferences across states and territories.

**Conclusion:** This study provides a globally unique insight into IgRT supply and demand in CLL patients by analysis of total national use in Australia over a six year period.

1. What is the NEW aspect of your work? (ONE sentence) The first and only comprehensive analysis of total national immunoglobulin replacement therapy (IgRT) facts and data of supply, demand and patterns of utilisation in Chronic Lymphocytic Leukaemia (CLL).
2. What is the CENTRAL finding of your work? (ONE sentence) Utilisation of IgRT (in this study period solely as IVIg) reflects the demographics of CLL patients with more advanced disease with hypogammaglobulinaemia and recurrent / severe infection (and hence in need of IgRT for infection prophylaxis), but utilisation can vary considerably.
3. What is (or could be) the SPECIFIC clinical relevance of your work? (ONE sentence) Immune failure occurs in virtually all patients with CLL and the resulting increase in infection risk is a major contributor to morbidity and mortality; this totally comprehensive, national data set over 6 years of IgRT use will inform all Haematologists (especially CLL clinicians and those managing patients with immunocompromise), State and National Blood Banks, State and National Departments of Health, immunoglobulin manufacturers and supply chains, and CLL and immunoglobulin replacement guideline authors in their understanding of IgRT supply, demand and utilisation.

## INTRODUCTION

Chronic Lymphocytic Leukaemia (CLL) is the most common low-grade B-cell lymphoproliferative neoplasm (1) and is invariably accompanied by some degree of immune impairment. Infective complications remain one of the leading causes of death and account for 50-60% of CLL-related mortality (2-11). Multiple patient and disease factors have been associated with the development of severe and/or recurrent (S&R) infections including increased age, advanced disease stage, co-morbidities, hypogammaglobulinaemia, a history of previous infections, and prior therapies. Many of these risk factors are not modifiable; however, several treatment modalities have been used to mitigate infection-related morbidity and mortality, including prophylactic antibiotics and the use of immunoglobulin replacement therapy (IgRT) in selected patients (12, 13).

Hypogammaglobulinaemia has long been recognised as an important risk factor for S&R infections in patients with both primary and secondary immune deficiencies since the 1960s (10). The literature reports wide variations in the incidence of patients with CLL developing hypogammaglobulinaemia with some series reporting it to be as high as 85.3% in patients with Binet Stage C disease (2, 3). Hypogammaglobulinaemia correlates with the frequency and severity of bacterial infections, particularly with encapsulated organisms such as *Streptococcus pneumoniae* and *Haemophilus influenzae*.

The use of regular IgRT, mainly intravenous immunoglobulin (IVIg), has been used to mitigate the incidence of S&R infections and sepsis-related mortality. During the 1980s and 1990s, several uncontrolled studies and three small randomised controlled studies were performed to assess the clinical impact of IVIg in CLL patients with acquired hypogammaglobulinaemia and S&R infections (12-14). The results of all three randomised controlled studies were promising with a nearly 50% reduction in documented bacterial infections, but no improvement in overall survival.

In 2008 the Cochrane Collaboration reviewed available data and identified 9 trials that investigated the impact of prophylactic IVIg in 459 patients with lymphoproliferative neoplasms, including CLL and multiple myeloma from multiple centres across Europe and the USA (15). Only the three trials mentioned above were included in the meta-analyses, which reported that prophylactic IVIg did not improve all-cause mortality in patients with CLL (RR: 1.36; 95% CI 0.58-3.19); however there was a significant reduction in clinically documented infections by 51% (RR: 0.41; 95% CI 0.39-0.61). The review commented on the high expense and the adverse side effects associated with IVIg suggesting that it should not be universally administrated to all patients with CLL (15). The trials included in the Cochrane review used a dosing schedule of 0.4g/kg every 3-4 weeks and the authors suggested an evaluation of the clinical impact of this treatment after 6-12 months to ensure that IVIg was only continued under appropriate circumstances. There is still no consensus on the optimal dose, or frequency of prophylactic IVIg in patients with acquired hypogammaglobulinaemia due to CLL or other lymphoproliferative neoplasms, and the literature remains dominated by surveys, commentaries and reviews (16-19). We herein document the total national utilisation of IgRT as IVIg in Australia for the 6 sequential years 2009-2013 and analyse patient demographics with published data together with patterns of national use.

IVIg is an expensive therapy costing between $AUS54-78 per gram and given the often protracted dosing regimens required for IgRT in haematological malignancies including CLL, the total annual cost for this therapeutic blood product is considerable (20, 21). In 2011-12, the Australian Government spent AUS$204.4 million on IVIg products; this equated to a little over 20% of the annual blood and blood product budget allocated to support a country with a population of 22.8 million people at the time (20). Therefore, good stewardship for this scarce and valuable commodity is essential.

## METHODS

### Intravenous Immunoglobulin Governance in Australia

In Australia, CSL Behring Australia is solely responsible for the manufacture of domestic IVIg (Intragam^®^ 10 introduced in 2017; Intragam^®^ P from 2000-2007 and Intragam^®^ prior to 2000) under a toll fractionation agreement with the Australian Government from the plasma of thousands of non-remunerated whole blood and plasmapheresis donors collected by the Australian Red Cross Lifeblood (Lifeblood) (20).

This arrangement has been insufficient to collect adequate plasma for fractionation to meet increasing clinical demand for IVIg for more than three decades. Hence, in 2004, the body responsible for the national supply of blood and blood products, the National Blood Authority (NBA) contracted international manufacturers of IVIg to supplement the domestic IVIg product to meet the growing needs of the nation. The NBA also developed national strategies to improve the governance of blood and blood product use which included the 2007 publication of the first edition of the *Criteria for the clinical use of intravenous immunoglobulin in Australia (The Criteria)* (22, 23). These guidelines were based on a systematic review of literature that supported the therapeutic use of IVIg for a range of neurological, immunological and haematological conditions requiring immune modulation or immune replacement that would be approved and therefore, funded by the NBA Agreement (23). *The Criteria* support the therapeutic replacement of IVIg for acquired hypogammaglobulinaemia secondary to haematological malignancies including CLL; this indication accounts for the highest proportion of annual IVIg distribution, and is hence the focus of this study.

### Criteria for the clinical use of intravenous immunoglobulin in Australia (The Criteria) for CLL patients with hypogammaglobulinaemia and severe and/or recurrent bacterial infections (23)

During the study period from 2008 to 2013, all clinical requests for IVIg were assessed against *The Criteria* for eligibility by clinical staff at Lifeblood prior to product release. The requesting medical practitioner was required to confirm the diagnosis of CLL, the presence of hypogammaglobulinaemia and that the patient had suffered from S&R bacterial infections. A single treatment dose based on the approved patient’s weight would be distributed to an approved health provider (AHP) for use by that specific patient. Ongoing IVIg supply was dependent upon at least annual review and documentation of ongoing clinical benefit.

Mandatory demographic data and basic disease characteristics were captured by Lifeblood databases including unique patient identifiers, age, gender, diagnosis and IgG level prior to IVIg therapy. Detailed CLL data such as stage, treatment status, haematological parameters and prognostic factors such as immunoglobulin gene mutation or cytogenetics were not required or recorded. The data for this study was retrospectively obtained from these databases after Ethics approval had been granted (Lifeblood Human Research Ethics Committee). Only the details of patients with CLL receiving IVIg for acquired hypogammaglobulinaemia and S&R bacterial infections were retrieved for inclusion in the study. The patient information was initially identifiable to allow the validity of the data to be assessed by the chief investigator and other key personnel named in the Ethics application and the Collaborative Research Agreement through secured computer networks to ensure the privacy of these patients was protected. All identifiable data obtained from these databases were destroyed following the completion of the study.

The time period selected for this study coincided with a change in the databases used at Lifeblood in 2008 through to 2013 after which Subcutaneous Immunoglobulin (SCIg) was approved for this indication in Australia. Virtually all national IgRT utilised by CLL patients with acquired hypogammaglobulinaemia and S&R bacterial infections was captured over this 6 year study period. Data from January and February 2008 was incomplete as some data was listed under an older classification system based on the AHMAC Blood and Blood Product Guidelines that was used to approve IVIg prior to the introduction of the first edition of the *The Criteria* in 2007 (21, 23). Therefore, when analysing annual data, only the 5 year data 2009-2013 were included.

## RESULTS

### Treatment Episodes

Lifeblood dispensed 48,870 treatment episodes of IVIg to 2,734 individual CLL patients, for acquired hypogammaglobulinaemia and S&R bacterial infections (hereafter referred to as “CLL”) across Australia from 2008 to 2013. The total volume of IVIg dispensed was 1,324,926 grams.

### Product Use

There were a total of six different IVIg products available to CLL patients during the study period. The vast majority of patients received the domestically manufactured Intragam^®^P (89.7%) with only small numbers of patients receiving imported products such as Octagam^®^ 5% and 10%, Kiovig10%, Flebogamma^®^ 5% DIF and Sandoglobulin^®^ NF (Table 1).

**Table 1.** IVIg Product Use from 2008 to 2013.

| Product | Origin | Treatment Episodes | % Use |
| --- | --- | --- | --- |
| Intragam® P | Domestic | 43845 | 89.72% |
| Octagam® 5% | Imported | 4603 | 9.42% |
| Kiovig 10% | Imported | 231 | 0.47% |
| Flebogamma® 5% | Imported | 111 | 0.23% |
| Octagam® 10% | Imported | 70 | 0.14% |
| Sandoglobulin® NF | Imported | 10 | 0.02% |

Legend: The six different IVIg products available to Australian CLL patients with acquired hypogammaglobulinaemia and S&R bacterial infections during the study period from 2008 to 2013.

### Patient Demographics

#### Age Distribution

Of the 2,734 CLL patients administered IVIg in the study period, the average age to receive their first treatment of IVIg was 74 years ±11 years. There were 2247 (82.2%) of patients over the age of 65 years, 836 (30.6%) between 65 to 74 years and 870 (31.8%) between 75 to 84 years (Figure 1). A small number of younger patients 35 (1.3%) received their first treatment of IVIg before the age of 44 years and a similar proportion 42 (1.5%) of very old patients received their first dose after the age of 95 years. This age data is consistent with Australian cancer registry data which reports the average age, at diagnosis (in contrast to age of commencement of IgRT), with CLL to be 69.9 years (24, 25) and comparable to international cancer registries including the USA, Europe and the United Kingdom (UK) where the median age of diagnosis is 70-72 years (26, 27).

**Figure 1.**
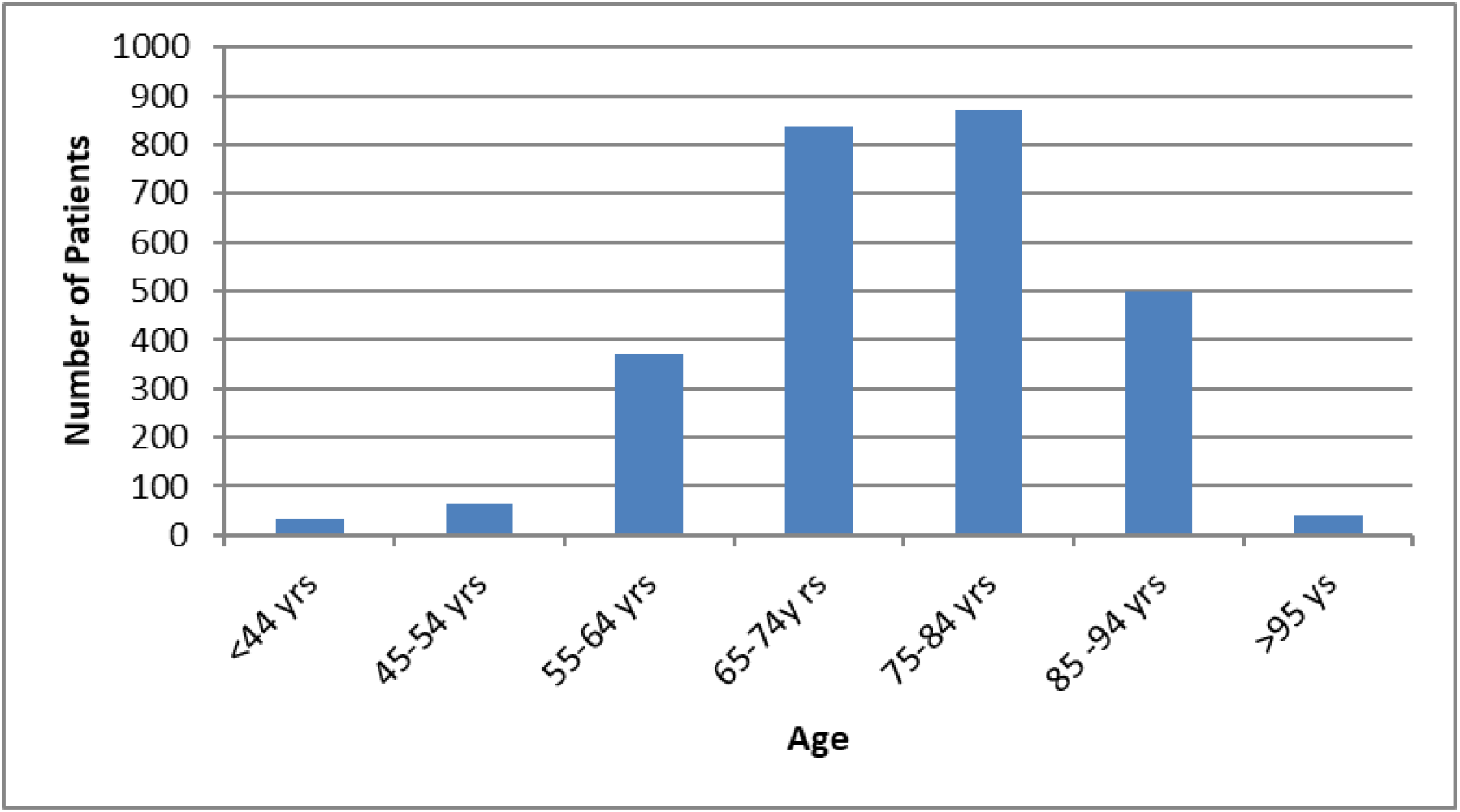
Age distribution of individual CLL patients. Age distribution of individual CLL patients based on when they received their first treatment episode of IVIg according to *The Criteria*.

#### Gender Distribution

Of the 2,734 individual CLL patients receiving IVIg, 59.3 % (1621 patients) were male and 40.7 % (1113) female. Overall male patients received 60.6% of the total treatment episodes representing 20% more treatment episodes of IVIg than female patients with CLL with cumulative utilisation by gender shown in Figure 2. Australian cancer registry data shows men are twice as likely to develop CLL with an age-adjusted incidence (ASI) of 6.0 per 100,000 males compared to females (M:F) with an ASI of 3.2 per 100,000 (24, 25). This gender ratio is comparable to the USA (M:F 6.75 and 3.65 per 100,000), Europe (M:F 5.87 and 4.01 per 100,000) (26), and the UK (M:F 8.1 and 4.8 per 100,000) (27).

#### IgG levels

The average pre-treatment IgG level was 4.03 g/L ± 2.03 g/L, with a wide range (0.30 g/L to 10.50 g/L). This was comparable to the average, baseline IgG levels of the 205 CLL patients included in the three trials reviewed by the Cochrane meta-analysis of 3.5 g/L ± 0.7g/L, 5.1 g/L ± 2.9 g/L and 4.8 g/L ± 2.9 g/L where the heterogeneity of the results were felt to be due to differences in the disease stages of the patients (15).

#### Progressive Annual Increase in Utilisation

Table 1 shows the total number of IVIg treatment episodes distributed by Lifeblood to patients with CLL over a five-year period (2009-2013). There was a sustained increase of 5.1% to 6.1% per year in the demand for IVIg as measured by increase in treatment episodes for these patients with an average annual growth rate of 5.5% (Figure 3).

**Figure 3:**
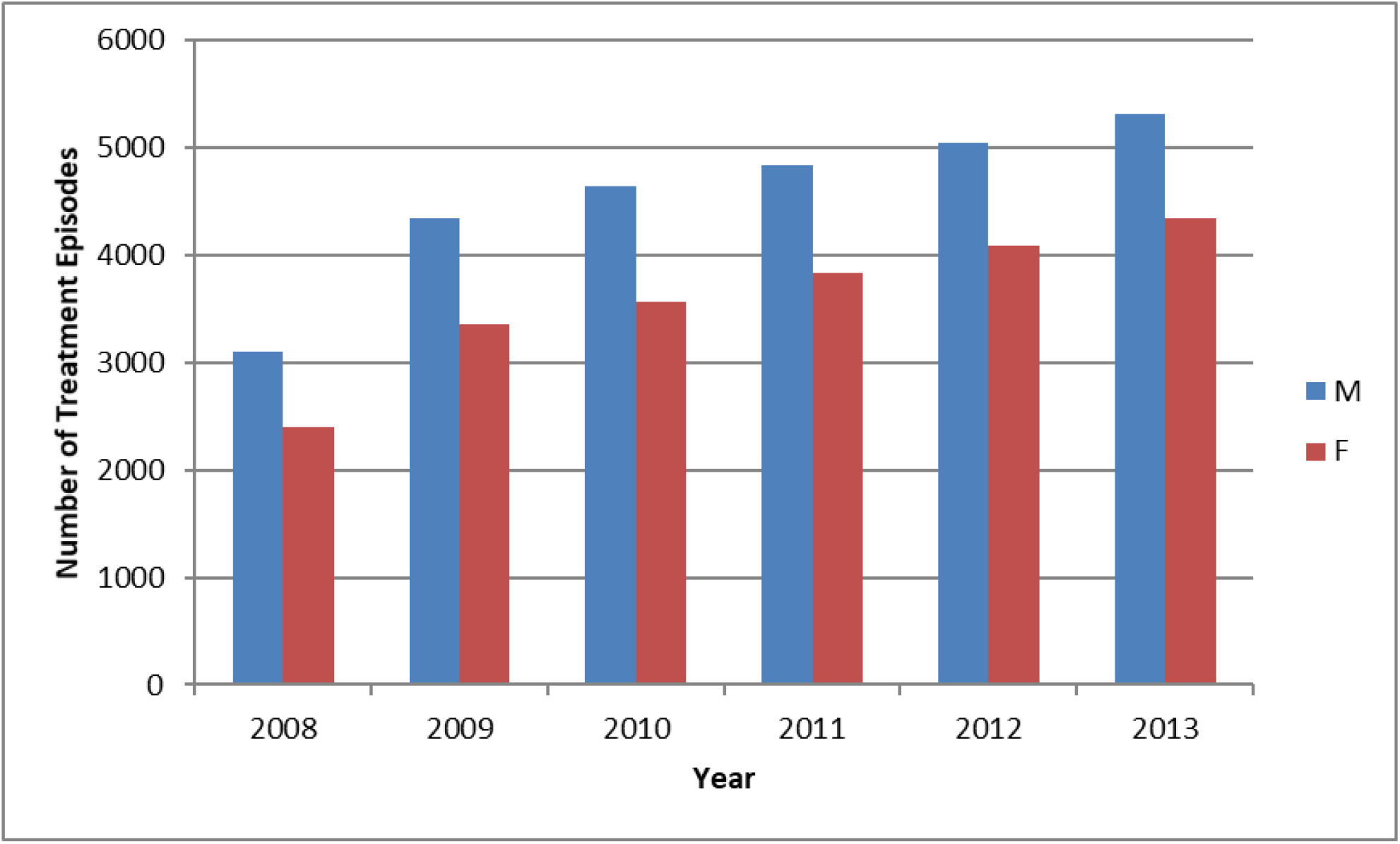
Cumulative IVIg utilisation by gender. Cumulative IVIg utilisation by gender from 2008 to 2013. M: male; F: female

#### Regional Australian State and Territory Utilisation

Examining IVIg distribution by number of treatment episodes (total 48,870), the majority of the national IVIg product was distributed to patients who resided in the three largest states, namely New South Wales (NSW) 16740 (34.3%), Queensland (Qld) 13851 (28.3%) and Victoria (Vic) 10244 (21.0%) (Table 2). Considerably less IVIg was distributed to the remaining smaller population three states South Australia 3766 (7.7%), Western Australia (WA) 1524 (3.1%) and Tasmania (Tas) 1524 (3.1%), and the two territories, Australian Capital Territory (ACT) 1095 (2.2%) and Northern Territory (NT) 126 (0.3%). Examining IVIg distribution by individual patients, the distribution of patients (total 2,734) largely matched number of treatment episodes and the three largest population states again had the highest numbers of individual patients as shown in Table 2.

**Table 2.** IVIg use by State and Territory, unique patients, and treatment episodes for 2011.

| **State** | **State/Territory**<br>**Population 2011**<br>**'000 (a)** | **Population %** | **Unique**<br>**Patients**<br>**per State**<br>**and**<br>**Territory** | **% IVIg by**<br>**unique**<br>**patients** | **% IVIg by**<br>**state &**<br>**territory**<br>**treatment**<br>**episodes** | **Treatment**<br>**episodes per**<br>**State and**<br>**Territory** | **Treatment**<br>**episodes per**<br>**1,000 capita** |
| --- | --- | --- | --- | --- | --- | --- | --- |
| **ACT** | **370.7** | 1.6% | 54 | 2.0 % | 2.2% | **1095** | **2.9** |
| **NSW** | **7,247.7** | 32.2% | 896 | 32.8% | 34.3% | **16740** | **2.3** |
| **NT** | **232.4** | 1.0% | 14 | 0.5% | 0.3% | **126** | **0.5** |
| **QLD** | **4,513.0** | 20.1% | 595 | 21.8% | 28.3% | **13851** | **3.1** |
| **SA** | **1,645.0** | 7.3% | 310 | 11.3% | 7.7% | **3766** | **2.3** |
| **TAS** | **511.7** | 2.3% | 86 | 3.1% | 3.1% | **1524** | **3.0** |
| **VIC** | **5,574.5** | 24.8% | 632 | 23.1% | 21.0% | **10244** | **1.8** |
| **WA** | **2,387.2** | 10.6% | 147 | 5.4% | 3.1% | **1524** | **0.6** |
| **Australian**<br>**Population**<br>**2011 (a)** | **22,485.3**<br>**in 2011 (a)** |  | **2734** |  |  | **48870** | **National**<br>**Average:**<br>**2.2** |

**Abbreviations:** ACT: Australian Capital Territory, NSW: New South Wales, NT: Northern Territory, QLD: Queensland, SA: South Australia, TAS: Tasmania, VIC: Victoria, WA: Western Australia. **Note** (a): Includes other Australian territories comprising Jervis Bay Territory, and Christmas, Cocos (Keeling) and Norfolk Islands. Australian 2011 population data reference: [3101.0 - Australian Demographic Statistics, Dec 2011 \(abs.gov.au\)](http://abs.gov.au/3101.0)

Analysis was undertaken to determine the proportion of IVIg distributed for CLL to each state and territory according to its population density based on 2011 population data (Table 2) (28, 29). This allowed direct comparison between the number of IVIg treatment episodes distributed to each state and territory per 1,000 capita. As shown in Table 2, during the study period, 32.2% of the Australian population lived in NSW and utilised 34.3% of the total IVIg which equated to 2.3 treatment episodes per 1,000 capita (Rx/1,000). Victoria, the second largest population (24.8%) used 21.0% of national IVIg (1.8 Rx/1,000) while Queensland with a smaller population of 4.47 million (20.1%) had the highest national rate of IVIg utilisation (28.3% = 3.1 Rx/1,000) across Australia. The two relatively small population Australian territories with comparable population, but dramatically different size demonstrated the differential utilisation most starkly. The Australian Capital Territory (ACT – total area 2,358 km^2^ and essentially only urban Canberra) with 1.6% of the Australian population had a utilisation rate of 2.9 Rx/1,000, the second highest rate of IVIg utilisation. By contrast, the Northern Territory (NT) with a large total area 1.42 million km^2^ with a population of 232,400 accounting for 1.0% of the Australian population (∼45% in the urban capital Darwin [2011 population 106,255], regional Alice Springs [2011 population 25,186] with the remaining 131,441 population widely dispersed in remote regional areas) had by far the lowest rate of IVIg utilisation of 0.5 Rx/1,000.

#### Monthly and Seasonal Distribution

Figure 4 graphically represents the monthly distribution of IVIg from 2009 to 2013 (the incomplete data from 2008 was not included in this trend analysis). There did not appear to be recurrent monthly peaks during the winter months of June, July or August as may have been expected due to the increased incidence of infectious complications during the winter season. There were sporadic unexplained peaks in the distribution of IVIg such as in October 2013. To analyse IVIg use by seasons of the year, the total monthly IVIg treatment episodes from 2009 to 2013 were divided into four seasons (summer, autumn, winter and spring). The cumulative use of IVIg appeared was lowest during summer months with 10363 treatment episodes (23.9%), increased slightly during the autumn months of 10835 (24.9%) and the winter months of 11020 (25.4%) with the highest use occurring in spring of 11150 (25.7%). The relatively minor reduction in the summer months coincides with the Christmas, New Year and typical January summer holiday periods during this season in Australia. The maximum variation of 1.8% between peak (spring) and trough (summer) seasons, with only 0.8% variation excluding the major holiday summer season, and higher use in spring than winter suggest that the practice of IgRT/IVIg use in winter with higher respiratory infection rates is not used to any significant extent in Australia.

**Figure 4.**
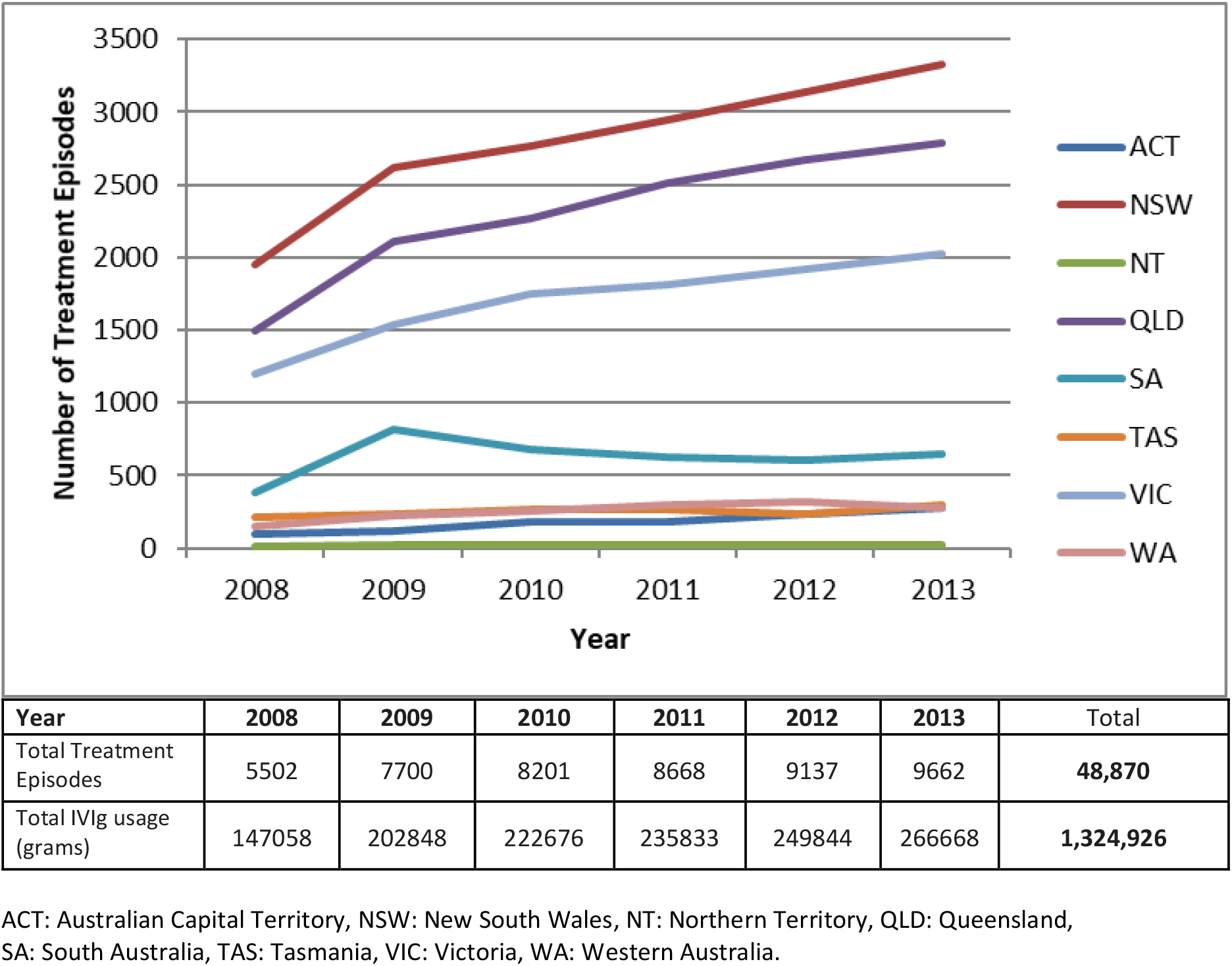
Trends in annual utilisation of IVIg for states and territories IVIg 2008 to 2013.

**Figure 5:**
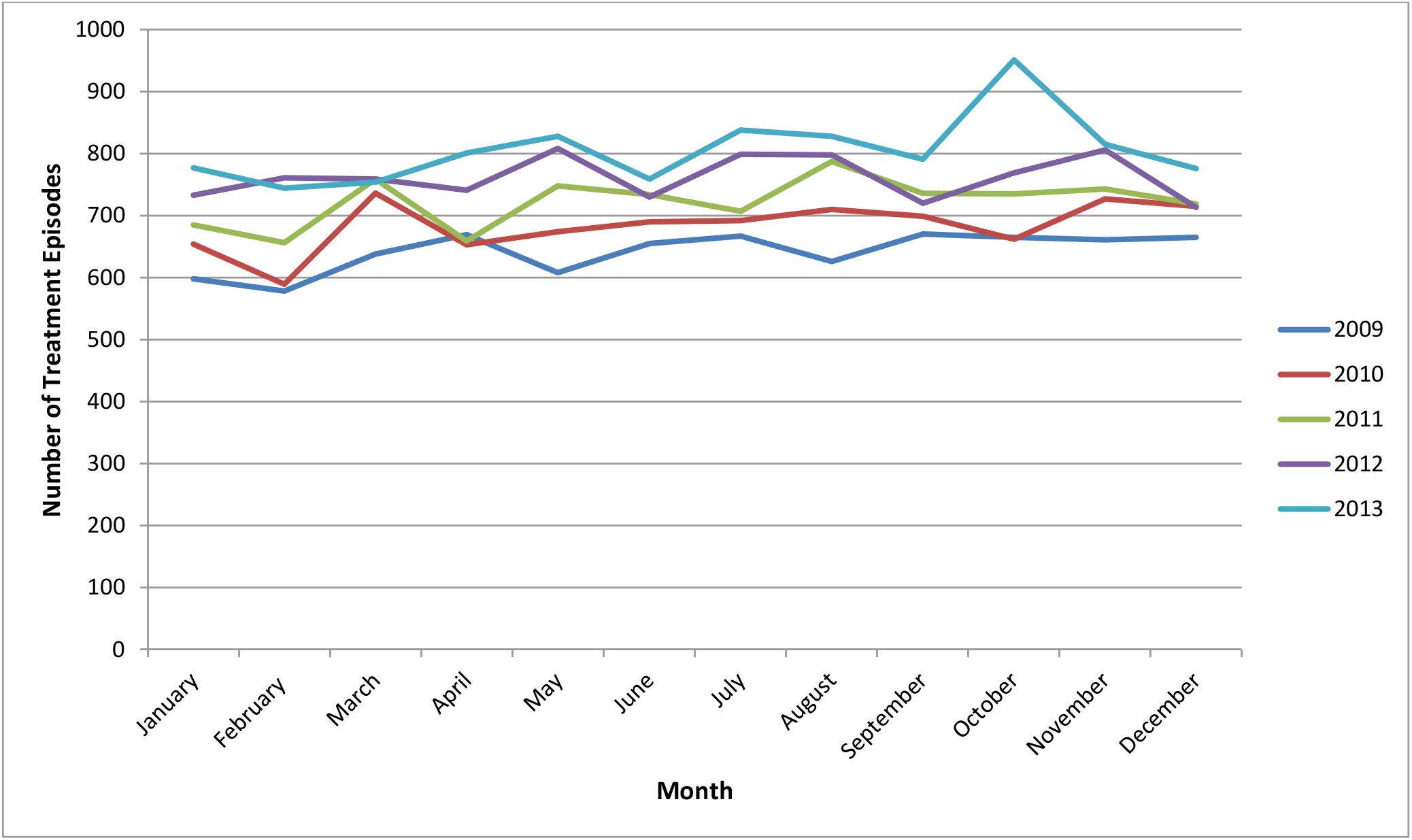
Seasonal variation by month for the years 2009-2013. Seasons by months in the Southern Hemisphere are summer: December to February; autumn: March to May; winter: June to August; and spring: September to November.

## DISCUSSION

In 2011-2012 (Australian Financial Year 1 July 2011 to 30 June 2012), total expenditure for finished IVIg products was $204.4 AUD million (approximately 22% of blood product expenditure under NBA arrangements, excluding the cost of collecting plasma for fractionation paid to the Blood Service) (23). Thus $204.4 million for 3.27 million grams of IVIg is an average cost of $62.51 per gram (noting domestic and imported IVIg different prices.). With 243,972 grams (∼ 6.6% of total IVIg) supplied for CLL patients in that year, the total IVIg cost for year 2011/12 for CLL patients was ∼$15.25 million, and hence on average $5,578.00 AUD per patient that year. To provide context, this is comparable with a single hospital admission via the Emergency Department in NSW ($7,557)(30), a single day in an Intensive Care Unit (ICU) bed (∼$4375) (31), or 6 months of fludarabine in 2011 ($6708) (32) and significantly less than adding rituximab to fludarabine plus cyclophosphamide (FC+R) in 2009 (incremental +R cost $40,268) (33). Furthermore by comparison, the cost of current targeted therapies is substantially higher again for both ibrutinib ($105,528 per annum) (34-37) and venetoclax ($93,410 per annum) (38, 39). Hence, the cost of ongoing IgRT/IVIg is offset if it prevents a single hospital admission via Emergency, or >1 day in an ICU bed each year.

Nevertheless, as patients with acquired hypogammaglobulinaemia secondary to haematological malignancies, including CLL, utilise the largest proportion of IVIg, it is vital for Australian, and international, healthcare sectors to understand the demographic characteristics and geographical distribution of patients to ensure adequate ongoing supply and good stewardship of this valuable resource. The demographic characteristics of the CLL patients in this study correspond with the epidemiological statistics from the Australian Institute of Health and Welfare (AIHW) for CLL including male predominance (∼2:1) and occurrence in the elderly, where the risk of CLL increases from 1 in 304 before the age of 75 years to 1 in 161 by the age of 85 years in Australia. The median age of diagnosis in Australia is 69.9 years and the median age for commencement of IVIg in this study is 74 years. This closely aligns with the typical time to commencement of therapy in CLL, ∼5 years after the diagnosis when both commencement of therapy (40) and some decline in immune function and IgG levels are seen, and also to the age of CLL patients starting IgRT in the German SIGNS study of 71 years (41).

One of the key findings of this study was that the national demand for IVIg for CLL patients with acquired hypogammaglobulinaemia and S&R bacterial infections continued to rise by an average of 5.5% per annum from 2008 to 2013, which resulted in Lifeblood and the NBA implementing strategies for increased plasma supply into the future. The 2011 Australian Census (29) reported an average population growth rate of 1.7%, and that the proportion of people over the age of 65 years had increased from 11.1% to 13.6% whilst the proportion over the age of 85 years had doubled from 0.9% to 1.8%. The largest growth rate of the very old was seen in the ACT (8.7%) followed by NSW and Victoria (both 6.3%), Queensland (6.2%), NT (6.0%), WA (5.9%), SA (5.4%) and Tasmania (4.7%) from 2009 to 2010. Hence, combined increases in population together with aging, and hence increasing numbers diagnosed with CLL, appear to account for much of the ongoing increase in demand in CLL, probably together with higher community expectations for standards of health care. This trend is likely to continue and will need to be factored into IgRT supply and budgets.

Anecdotal reports suggest that some clinicians utilise IgRT in a seasonal strategy. However, when we examined IVIg treatment episodes by seasons of the year, this was not the case, as more IVIg was distributed during spring than winter. Although summer was when the least number of treatment episodes of IVIg were distributed, this modest reduction corresponds with the Christmas, New Year and summer holiday periods in southern hemisphere Australia when hospital services and staffing are commonly reduced and this would appear to readily account for the 1.8% variation between peak and trough seasons. Hence, the strategy of seasonal use appears not to be a significant contributor to variation of use in Australia.

Not surprisingly, the three states with the largest populations, NSW, Queensland and Victoria, received the majority of the IVIg. Nearly 85% of national IVIg was distributed to these states compared to a combined population of 75% of the Australian population at that time. The NBA blood and blood product usage report for 2011-2012 suggested that Tasmania, Queensland, NSW and the ACT utilised the most IVIg across all indications approved by *The Criteria*. Analysis in this study confirmed that Queensland (3.1 Rx/1,000), Tasmania (3.0 Rx/1,000), the ACT (2.9 Rx/1,000), NSW (2.3 Rx/1,000) and SA (2.3 Rx/1,000) used more than the national average of 2.2 treatment episodes of IVIg per 1,000 capita. This finding was similar to the NBA report, although the order of the states and territories differed slightly. While there are differences in patient demographics between state and territories, individual physician and institutional prescribing practices appear to explain these findings.

Remoteness is an issue in Australia with a very large area yet a highly urbanised population. Evaluation of remoteness has been defined (42-44) for health and other services and in 2009 there was 69.2% of the population in ‘major cities’, 19.4% in ‘inner regional areas’, 9.1% in ‘outer regional’, while 1.5% and 0.8% lived in ‘remote’ and ‘very remote’ areas respectively. Hence while remoteness will likely have had an impact on IVIg access for a small number of individual patients, it is insufficient to explain state and territory differences. For patients in remote areas, self-administration of subcutaenous Ig (SCIg) since 2013 should partially address the issue of remoteness and access. Age data from the 2011 Census (ABS) showed Tasmania had the oldest population of all the states and territories in 2011 (median 40.2 years) and the greatest increase in its mean age from 1990 to 2010 which may have contributed to the second highest IVIg use per capita. Queensland had the highest national rate of IVIg per 1,000 capita for CLL, however the proportion of people over the age of 65 years in the three most heavily populated states, namely Queensland, NSW and Victoria, are consistent with the national average suggesting that the discrepancy between Queensland’s population (20.1%) and the proportion of national IVIg supply (28.3%) was due to the prescribing preferences of individual physicians who care for CLL patients. Furthermore, the striking difference of IVIg use between the two territories (ACT – population 370,700 and NT population 232,400) is more in keeping with prescribing practice rather than remoteness and age. Finally, Victorian use is lower (1.8 Rx/1,000) than the other large population states of NSW and Queensland despite virtually the entire population living within 2 hours of a capital or regional hospital, similarly consistent with lower precribing rates rather than age (median age NSW 37.4 years, Victoria 37.1 years, Queensland 36.4 years in 2011) or access as the primary factor determining utilisation.

IVIg is a relatively scarce and valuable blood product used to treat a range of clinical conditions, including IgRT for CLL patients with hypogammaglobulinaemia and S&R bacterial infections. The estimated cost while not insignificant is offset if it prevents one hospital admission per year and is comparable to other forms of CLL therapy (11), and significantly less than targeted therapies. There was a steady annual increase in demand for IVIg for this indication in Australia from 2008 to 2013. This 6 year time frame uniquely captured all IgRT utilisation for CLL across the entire country following the implementation of new national guidelines under the governance of a single institution, the Australian Red Cross Lifeblood. The demographics of IgRT utilisation closely reflected the demographics of CLL in Australia. The study has demonstrated significant variations in the distribution of IVIg between states and territories. Trend analysis of population demographics including age and gender together with monthly, seasonal and yearly IVIg distribution, and remoteness explain some of these variations. However, the impact of the prescribing preferences of individual physicians and institutions who manage CLL patients appears to be the most significant influence on the demand for IVIg.

These results provide valuable demographic and distribution data about IgRT in CLL for future clinical management and provide insights for all Haematologists (especially CLL clinicians and those managing patients with immunocompromise), the NBA Australia and other state and national blood banks, state and national Departments of Health, immunoglobulin manufacturers and supply chains, and CLL and immunoglobulin replacement guideline authors, and other stakeholders in their understanding of IgRT supply, demand and utilisation.

## Data Availability

All data produced in the present work are contained in the manuscript

## Acknowledgement

Australian governments fund Australian Red Cross Lifeblood to provide blood, blood products and services. Thanks to Dr Yandong Shen for assistance with referencing.

